# Effect of implementing population-based prostate-specific antigen screening on testing rates and prostate cancer overdiagnosis in England: a statistical modelling study

**DOI:** 10.64898/2026.01.23.26344710

**Authors:** Andrew J. Vickers, Veeru Kasivisvanathan, Adam Brentnall

**Author notes:** **Corresponding author:** Andrew Vickers, Memorial Sloan Kettering Cancer Center, 1275 York Avenue, NY, NY 11215.

## Abstract

**Objective:** To determine the potential effect of implementing population-based prostate-specific antigen (PSA) screening in England on overdiagnosis and testing rates compared with the current opportunistic testing policy.

**Design:** Statistical modeling study. English data on rates of prostate cancer by stage, PSA testing, and life expectancy were merged with epidemiological data on lead time to evaluate plausible overdiagnosis and PSA testing rates from an organized population-based program, in comparison with the current opportunistic policy. In the base-case scenario, organized screening increased the rate of PSA testing in men aged 50 – 69 year and decreased PSA testing in older men. An alternative modeling approach estimated change in overdiagnosis using data from the UK “CAP” trial.

**Setting:** England, 2018/19.

**Participants:** Adult men.

**Main outcome measures:** Rates of PSA testing, early-stage prostate cancer incidence, and overdiagnosis (prostate cancer that would not be diagnosed in a man’s lifetime but for the PSA test).

**Results:** In the base scenario, introduction of population-based screening led to an approximate 25% reduction in both PSA testing and overdiagnosis rates in the target population compared with the current policy. This was due to the anticipated decrease in PSA testing and overdiagnosis in men aged 70+ years being larger than the projected increase in PSA testing and overdiagnosis in men 50-69 years. The overall incidence of early-stage cancer prostate cancer was similar. Population-based screening was found to detect more early-stage cancers that were not overdiagnosed, and is therefore likely to have a greater impact on prostate-cancer morbidity and mortality than current policy. Findings were robust in sensitivity analyses including the entirely independent modeling approach based on CAP.

**Conclusion:** Opportunistic screening policies in England have led to high rates of overdiagnosis and PSA testing. In comparison with current policy, a risk-adapted, population-based prostate cancer screening program would likely reduce the number of PSA tests and overdiagnoses, and increase benefits of PSA testing from reduced prostate-cancer mortality Population health in England would be improved by either adopting an organized program or by ending PSA testing in primary care.

**Research in context:** *Evidence before this study:* It is known that screening for prostate cancer using the PSA test results in considerable overdiagnosis. As a result of these concerns, UK national policy, like most high-income countries, is to have no population-based screening program but to allow men to obtain a PSA test after shared decision-making with a primary care physician. This has led to very high rates of PSA testing among men aged over 70, who experience limited benefit from screening but who are at high risk of overdiagnosis. A small number of studies have modeled the impact of implementing population-based PSA screening but have not incorporated how such a program would affect opportunistic screening in older men.

*Added value of this study:* Data on rates of prostate cancer by stage, PSA testing and life expectancy in England were merged with epidemiological data on lead time to model statistical rates of overdiagnosis and PSA testing from an organized population-based program in comparison to current policy. Changes in PSA testing in men above screening age were based on how empirical data on how rates changed in the one country that has implemented population-based screening, Lithuania. We estimate that introduction of population-based screening would reduce both PSA testing and overdiagnosis by about 25%. This is due to the anticipated decrease in PSA testing and overdiagnosis in men aged 70+ years being larger than the projected increase in PSA testing and overdiagnosis in men 50-69 years subject to the population-based screening program. Population-based screening was found to detect more early-stage cancers that were not overdiagnosed and is therefore likely to have a greater impact on prostate-cancer morbidity and mortality than current policy. Findings were robust in sensitivity analyses including an entirely independent modeling approach.

*Implications of all the available evidence:* Opportunistic screening policies in England have led to high rates of overdiagnosis and PSA testing. In comparison with current policy, a risk-adapted, population-based prostate cancer screening program would likely reduce the number of PSA tests and overdiagnoses, and increase benefits of PSA testing from reduced prostate-cancer mortality Population health in England would be improved by either adopting an organized PSA screening program or by ending PSA testing in primary care

## Introduction

Screening for prostate cancer using prostate-specific antigen (PSA) is not currently recommended by the UK National Screening Committee due to “concerns about high levels of overdiagnosis [and] overtreatment”.^1^ Accordingly, there is no organized National Health Service (NHS) PSA screening program comparable to the NHS Breast Screening Program with mammography, or the NHS Bowel Program with fecal immunochemical tests. In both these programs, the target population is proactively contacted at a determined interval, provided with information for informed decision-making, and if desired, invited to take the screening test.

In place of population-based screening, the current NHS policy is known as the “Prostate Cancer Risk Management Program”. This is an opportunistic testing program^2^ whereby “any man can make an appointment with their GP to discuss having the … PSA test” ^3^ and, following such informed decision-making, “anyone aged 50 or over with a prostate can ask for a PSA test”^4^. Moreover, as part of national guidelines on recognition of suspected cancer^5^, a PSA test is recommended for men with lower urinary tract symptoms (LUTS), erectile dysfunction or hematuria. As a result, many men in the UK receive PSA tests. Collins et al. reported data on the incidence of PSA testing in England in 2018, finding that among ∼10m men followed for a mean of ∼8 years, 1.5m had close to 4m PSA tests^6^. They also noted that PSA testing was much higher for men in their 70s, 80s and 90s compared to those aged 50 – 70 years. Overdiagnosis of prostate cancer is strongly associated with age^7^ and therefore opportunistic testing policies have led to very high rates of PSA screening in the groups of older men who are at highest risk for overdiagnosis^8^.

Given that the current UK PSA testing policy results in overdiagnosis, a relevant question for policymakers is how the introduction of population-based PSA would change rates of overdiagnosis in the population compared with the current *status quo*. By population-based screening we mean a program where PSA testing is organized by the NHS through a call and recall system, following a defined screening regimen that includes screening intervals, triage tests, diagnostic testing, referral and appropriate information technology systems for monitoring. PSA testing outside of the program would be restricted to urologists and not be available in primary care. To estimate of how rates of PSA testing and overdiagnosis would accordingly be expected to change with such a program, we undertook a statistical modeling study merging data on current rates of PSA testing and prostate cancer diagnoses by stage with predicted changes in screening rates by age group that might be expected following the introduction of population-based PSA testing.

## Methods

An overview of the data sources and scenarios used in the base case model is given in Table 1. National statistics were obtained on the number prostate cancer cases in England annually, and the proportion of cases that are clinical T-stage 1 (T1) and T2 versus T3 and T4, separately by age group (less than 50, 50 - 59, 60–69, 70–79 and 80+ years). Our base case scenario used rates from 2019, to match PSA testing data from Collins et al.^6^

**Table 1.** Overview of data sources and scenarios for each model input. All inputs are available in the Stata code included in the Supplementary Material.

| Input | Data source | How varied in sensitivity analysis | Comments |
| --- | --- | --- | --- |
| Number of early and late stage prostate cancers diagnosed by age group | NHS data<sup>20</sup> | Not varied |  |
| Life expectancy by age group | UK life tables<sup>21</sup> | Not varied |  |
| Male population by age | UK Office of National Statistics<sup>22</sup> | Not varied | Data available for England and Wales. Assumed 50% male and took into account relative population in England vs. Wales and size of population 90+ vs. 80 – 90. |
| Current PSA testing rates by age group | Study on UK PSA testing<sup>6</sup> | Not varied |  |
| Proportion of PSA tests given for symptoms of prostate cancer | Study on UK PSA testing<sup>6</sup> | Varied the probability that existence of prior symptoms led to PSA testing from 100% to 10% with 80% as the base case. | Insensitive to changing assumptions, with ~2% difference between 100% vs. 10% rates. |
| Proportion of prostate cancers diagnosed after “for cause” biopsy | UK primary care audit study<sup>23</sup> | Varied the probability that existence of prior symptoms led to biopsy from 100% to 10% with 80% as the base case; also varied how this probability differed between early and late stage cancers from odds ratio of 1 for late stage cancer (i.e. no difference) to an odds ratio of 3 for late stage cancer with 2 being the base case | Estimates highly sensitive to rate of “for cause” cancers, discussed further in text. |
| Compliance | UK compliance with bowel and breast screening both ~70%<sup>24</sup> | Varied from 50% to 100%, with 70% as the base case | Moderately sensitive to changing assumptions. Under base scenario, varied from 69% relative number of diagnoses with 50% compliance to 85% with 100% compliance |
| Lead time | ERPSC<sup>25</sup> | Varied from 5 to 15 years, with 7 years as the base case | Relatively insensitive to changing assumptions, with less than 6% difference in relative overdiagnosis rates between 5 vs. 15 year lead time. |
| Change in PSA use in older men with population-based screening | Lithuania study<sup>10</sup> | Varied from 80% (base case) to 50% | Sensitive to changing assumptions, with a 50% vs.80% reduction in screening leading to roughly half the reduction in overdiagnosis associated with population-based screening. |
| Proportion of men overdiagnosed irrespective of life expectancy | ERPSC<sup>26</sup> | Varied from 0 (base case) to 20% | Number of patients overdiagnosed changes, but relative number between current practice and population-based screening unaffected. |

Our mathematical model is described in detail in the Supplementary Appendix. In brief, we took overdiagnosis to be negligible in men diagnosed with stage T3 and T4 disease. The risk of overdiagnosis for T1 and T2 prostate cancer for each decade of age was calculated in terms of the lead time and life expectancy and then multiplied by the number of men undergoing PSA testing in that age group to calculate total number of overdiagnoses. Changes in overdiagnosis with the implementation of population-based screening were estimated by increasing the number of men aged 50 – 69 undergoing PSA testing (to 70% in the base case, comparable to compliance with other cancer screening programs^9^) and reducing PSA testing in older men (by 80% in the base case, what has been empirically observed in older men follwing the implementation of a population-based PSA testing program^10^). Current UK policy is that a PSA should be given if requested by a patient or “for cause”, if a patient reports certain symptoms, such as erectile dysfunction. We separately modeled changing just the former or both of these policies, in other words, population-based screening would be implemented with a policy that men could not request PSA, with or without a separate policy that a PSA could not be ordered in primary care in response to symptoms. It is of note that T1 and T2 prostate cancer does not cause symptoms and therefore the most rational policy is to have no “for cause” PSA testing primary care. To calculate the number of PSA tests under population-based screening, we followed the algorithm suggested by European guidelines as to the frequency of PSA testing at different ages^11^ We triangulated our findings using an independent modeling strategy. This was derived using the cumulative incidence after screen detection from a screen using 15 years of follow-up from the UK CAP trial of PSA testing^12^. Code for both approaches is given in the Supplementary Material allowing independent researchers to change the inputs to determine how it affects the findings.

## Results

The results for the base case scenario are shown in Table 2. The number of PSA tests under population-based screening was reduced by approximately 25% compared with current policy. This was driven by a very large decrease, from >900,000 to <200,000 men aged 70 years or older, which more than offset the approximate expected doubling of PSA testing in men in their 50s. The number of PSA tests in men in their 60’s was also reduced, as the population-based strategy excluded from screening men with PSAs < 1 ng/ ml, constituting about half of this age group.

**Table 2.**
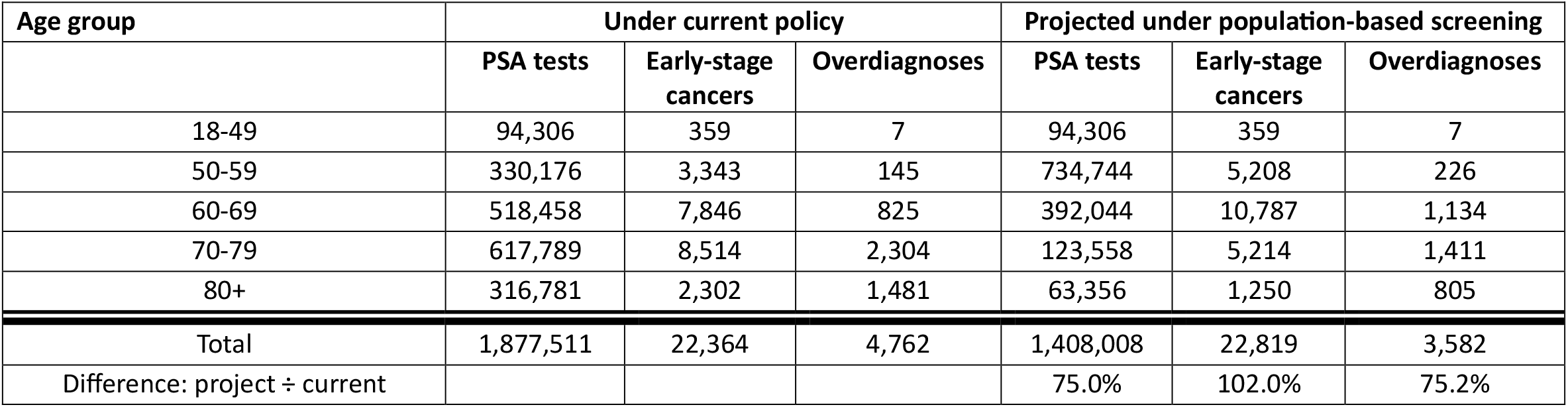
Estimated PSA tests, early-stage cancers (stage T1 or T2) and overdiagnoses under current policy and projected under population-based screening for the base case scenario.

The projected effect of population-based screening on overdiagnosis was also close to a 25% reduction, again driven by a large decrease of early-stage cancers in older men. The absolute number of overdiagnosed men did not increase by a large amount in younger men, in line with the model assumption on the longer life expectancy of younger men, and hence greater likelihood that a cancer would eventually be clinically diagnosed during their lifetime. The anticipated overall number of early-stage cancers was very similar under population-based screening, with increased incidence in younger men offset by a decrease in older men. There was a ∼10% increase in the number of early-stage cancers that are not overdiagnosed.

Table 3 shows results from the sensitivity analyses. Given that the base case showed an advantage to population-based screening, we focused on changes that would be expected to reverse this result. However, even with extreme assumptions, such as 100% compliance with population-based screening, no change in “for cause” biopsies and only a 50% reduction in screening in older men, the number of men overdiagnosed did not importantly increase with population-based screening. We examined one further scenario, which was a more aggressive approach to reducing “for cause” PSA testing and biopsy. This reduced overdiagnosis by about half compared to contemporary policies, driven by a large decrease in early-stage cancers in men aged 70 and over.

**Table 3.** Results of sensitivity analyses changing various inputs and assumptions.

| **Scenario** | **Relative number of overdiagnoses with population-based screening compared to current policy** |
| --- | --- |
| Base case | 75.2% |
| Alternative scenario 1. Symptoms before biopsy were always cause of biopsy and no difference in symptomatic presentation between early and late-stage cancers | 93.9% |
| Alternative scenario 2. Compliance rate with population-based screening 100% in men of screening age | 84.6% |
| Alternative scenario 3. Decrease in screening older men only 50% with population-based screening | 87.6% |
| Combine scenarios 1 – 3 | 102.5% |
| Base case with additional policy that PSA cannot be ordered in primary care in response to symptoms such as hematuria | 47.5% |

The results of the independent analysis using the CAP data are shown in figures 1 and 2. The results are consistent with the primary analysis. In the base scenario for overdiagnosis (figure 1a), if introduction of population-based screening led to an 80% reduction in screening in men 70 and greater screening and an increase in overdiagnosis in younger men of about 50%, there would be approximately 1000 fewer overdiagnoses than currently, very similar to the estimate of around 1200 shown in the primary analysis (table 2). Figure 2 shows that if population-based screening reduces incidence of prostate cancer in men older than 70 years by 80%, detection would need to increase by more than 250% in men aged 50-69 to increase overdiagnosis relative to the current policy. This is impossible given that close about 40% of men aged 50 – 69 already received PSA tests.

**Figure 1:**
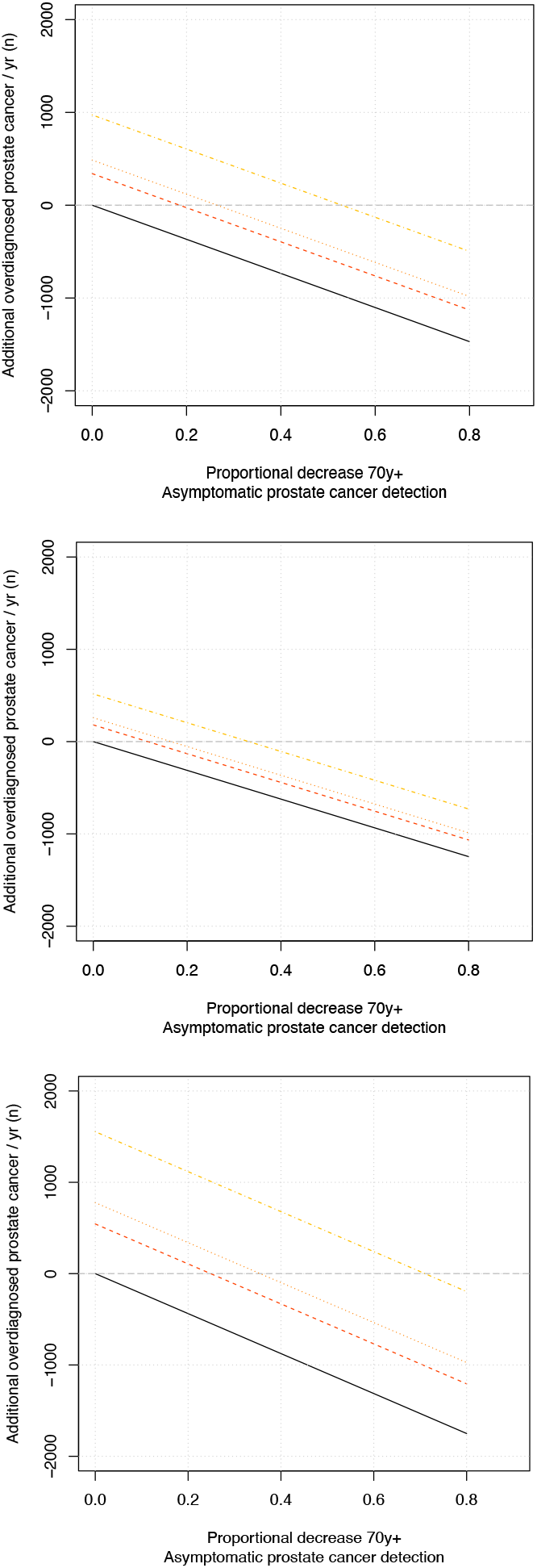
Change in estimated number of overdiagnosed prostate cancers detected each year in England from population screening using data from CAP. Change in detection of cancers in men aged 50-69 years is given as a factor of 1 (ie. no increase (black) 1.35 (red), 1.5 (orange) or 2 (yellow). Plot (a) shows results under the base scenario for overdiagnosis from the CAP trial; (b) shows results based on the lower bound of the 95% CI for overdiagnosis from the CAP trial; (c) is based on the upper bound of the 95% CI.

**Figure 2:**
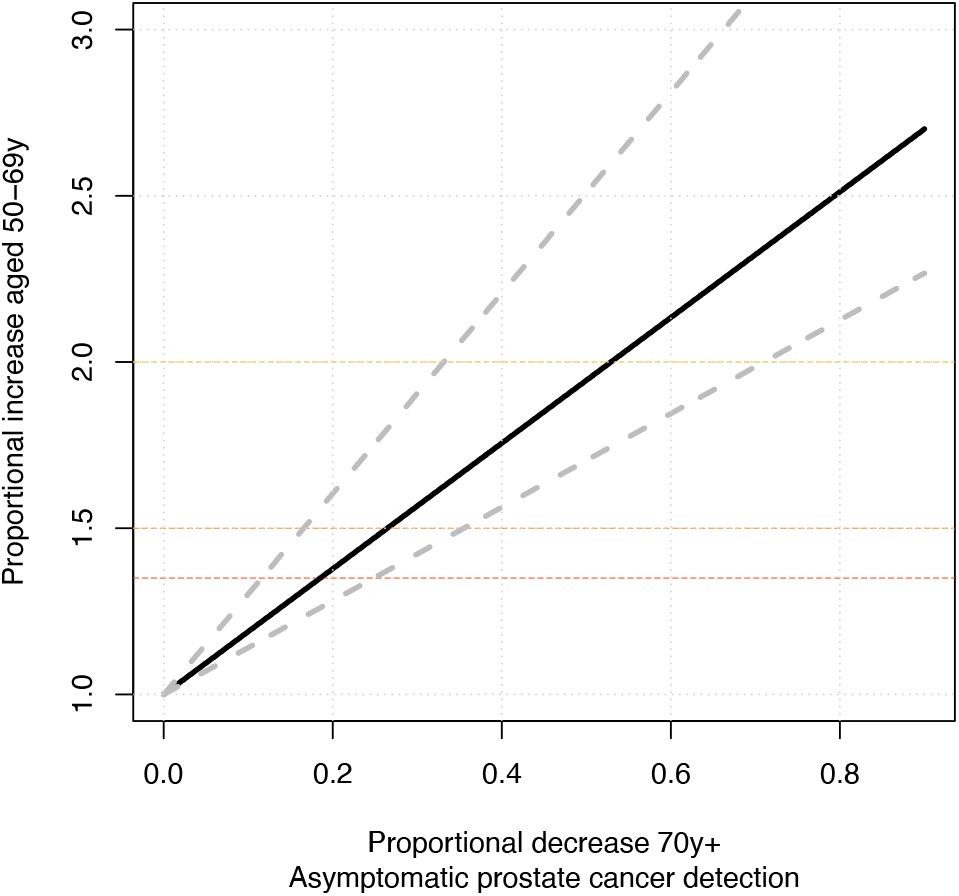
Required change of detection in older men to reduce overdiagnosis, using CAP data. Estimated proportional decrease in detection rate of cancer from population-based screening compared to current opportunistic policy required in men aged 70+ required to match the increase in overdiagnosis in men of screening age. The base scenario estimate (black) is shown with estimates from the upper and lower 95% CI overdiagnosis estimates (gray).

## Discussion

Using a simple modeling strategy, we found that, in comparison with the current opportunistic testing policy, implementation of population-based PSA screening in the UK is unlikely to substantially increase either the number of prostate cancer overdiagnoses, or the number of PSA tests done annually. Provided that access to PSA testing is only available through the screening program, or from a urologist, the anticipated increase in PSA tests and overdiagnosis associated with a roll out of population-based screening in younger men is likely to be offset by a countervailing decrease in older men when PSA tests were no longer available in primary care. In the base model scenario, population-based screening was expected to reduce both PSA testing and overdiagnosis by about 25%. In another reasonable scenario, abandoning the questionable policy of recommending PSA testing in primary care for symptoms such as erectile dysfunction, hematuria and LUTS, the total incidence of overdiagnosis was approximately halved.

Screening for prostate cancer using PSA testing leads to overdiagnosis, and so it might appear counterintuitive that implementing a program to provide PSA tests would reduce overdiagnosis. However, it is perhaps unsurprising that a rational program specifically designed to reduce overdiagnosis would have better outcomes than a *laissez-faire* approach where a man can get a PSA if he asks for one, even if he has too short a life expectancy to benefit.

Our findings also suggest that implementation of population-based screening would provide greater benefit than current policy. This is because the number of T1 and T2 cancers that are not overdiagnoses increases by about 10%. Such cases represent cancers detected early and thus represent a chance of cure of a disease that might otherwise progress to metastasis.

It is sometimes argued that implementation of population-based screening would require considerable extra resources to cope with the increase in cases and would thus place undue strain on an already over-burdened health system.^13^ Our findings suggest this would not be the case in the UK. Provided that increased detection in younger men is accompanied by reduced detection in older men, population-based screening was found unlikely to increase either the number of PSA tests done annually, nor the total current number of early-stage prostate cancers detected, in comparison with the current *status quo*. Further, one might also anticipate a reduction in the number of advanced stage cancers due to the increase in early-stage cancers that are not overdiagnosed. Given the very high resource requirements for advanced disease, this suggests that implementation of population-based screening might also result in a decrease in resource requirements for prostate cancer. We also postulate that population-based screening would alleviate burden on primary care physicians, who are asked to engage in complex and lengthy shared decision-making with patients about a subject in which their expertise may be less than complete. For instance, one shared decision-making proposal is that GPs inform patients on 16 separate facts and ask 12 questions about preferences^14^.

Other possible advantages of organized screening over opportunistic testing that are beyond the scope of our analysis. One of these is that it should reduce the major well-documented regional and income disparities associated with the opportunistic testing. In the Collins et al. analysis of PSA testing, for instance, rates of PSA testing in the north-east of England were over 40% lower than in the south-east; there was a similar difference in PSA testing comparing the highest versus lowest quintile of deprivation^6^. Dodkins et al. have demonstrated how such variations lead to differences in rates of metastatic prostate cancer^15^, with an approximate relative 10% increased risk comparing the least to highest deprivation quintile.

Our estimates of the risk of overdiagnosis are in line with those reported elsewhere in the literature. In perhaps the most well-known study^16^, the risk of overdiagnosis for systematic biopsy following an elevated PSA was estimated as “23% to 42% of all screen-detected cancers”. The use of MRI to determine biopsy, which is very widespread in the UK[41392198], reduces diagnoses following a PSA test by about 40%[PMID 39321360]. Our estimate of 4,762 from 22,364 (21%) cancers overdiagnosed is therefore approximately in the middle of the expected range. Our estimates showing reductions in overdiagnosis of 25 – 50% with restriction of screening in older men also have prior support in the literature. A comparison of prostate cancer incidence data before and after the introduction of PSA screening in the US found that 42% of the excess cases attributable to PSA were in men aged 70 and older.^7^

One critical input to our model is the reduction in PSA testing in older men with the implementation of organized screening. Our base case estimate of an 80% reduction might be considered unrealistic, but it is critical to note that this is reflects anticipated policy not predicted behavior: it is not that we expect older men to stop asking for PSA tests, or GPs to stop ordering them, simply because a screening program is in place; our scenario assumes instread the NHS will make policies to make PSA testing rarer in older men, for instance, by preventing PSA testing in primary care outside defined indications, such prior prostate cancer. This is exactly what happened in Lithuania, where the observed 80% reduction in screening in older men occurred following changes in coding, billing and budgeting policies for PSA testing in primary care.

Two other models have been built to evaluate the potential utility of organized prostate cancer screening the UK, but one of the motivations for this work was an observation that neither do not explicitly consider the level of PSA testing ongoing in the UK, nor model how formal screening would reduce opportunistic PSA testing in older men. For example, one report on how a national prostate cancer screening program would affect NHS costs and activity^17^ found that a population-based screening would increase the number of prostate cancer diagnoses. This is because the model focused only on men aged 45 – 69 and assumed no change in PSA rates in men aged 70 and above. Another model^18^ similarly did not incorporate how implementation of population-based screening would affect rates of opportunistic screening, especially in older men and, thereby also implicitly assumed that screening would be an “add on” to existing activity. We do not believe “population-based screening plus PSA testing whenever a man wants it” is a reasonable policy and therefore do not believe estimates of the effects of such a policy are of relevance to decisions about PSA policy.

A possible alternative to both the current informed choice policy and population-based screening would be to maintain the current *status quo* but restrict PSA testing both in older men and in response to symptoms. For instance, the current policy might be amended from “anyone aged 50 or over with a prostate can ask for a PSA test” to “anyone aged 50 - 69 with a prostate can ask for a PSA test”; guidelines might also be updated to remove the recommendation for PSA in men with LUTS, erectile dysfunction or hematuria. While this would reduce the high level of overdiagnosis under the current policy, it would not address the policy problem of regional and economic disparities.

Our simplified modeling approach is subject to several limitations. For instance, we did not take into account the effect of repeated PSA tests, and assumed that diagnoses occurring within an age group (e.g. 70 – 79) took place at the midpoint age (e.g. 75). Critically, however, there is no reason to believe that such limitations would importantly bias our findings towards population-based screening. In particular, any such bias would have to be extremely large to change our main conclusion that the introduction of population-based screening would not increase prostate cancer overdiagnosis.

In conclusion, implementation of an organized, population-based prostate screening program would not importantly increase PSA testing and overdiagnosis in comparison with current NHS policies and guidelines on PSA testing. Indeed, such a program would most likely reduce the harms and costs of screening as well as increase benefits in terms of mortality. As such, our findings suggest that current NHS policies and guidelines on PSA should be changed. The NHS should “fish or cut bait”^19^, either prohibit the use of PSA in primary care or implement a comprehensive, risk-adapted, population-based screening program designed in response to best evidence on how to reduce the harms and maximize the benefits of PSA screening.

## Data Availability

All data produced in the present work are contained in the supplementary material

