## Supplementary material for "Effect of implementing population-based prostate-specific antigen screening on testing rates and prostate cancer overdiagnosis in England: a statistical modelling study"

**Supplementary appendix**

**Methods**

We assumed that any man with T3 or T4 cancer would have been diagnosed during the course of his life irrespective of PSA. To estimate annual rates of screening-related overdiagnosis in patients with T1 and T2 prostate cancer, we considered that a proportion of T1 and T2 would be related to work-up for symptoms of prostate cancer. Our inputs for this parameter used data from the 2018 National Cancer Diagnosis Audit^1^ which reported the proportion of patients diagnosed with prostate cancer after asymptomatic PSA testing, separately by age group. Patients were defined as having asymptomatic PSA only if there was no prior record of the symptoms defined in NHS policy to require a PSA (LUTS, erectile dysfunction, hematuria). However, the presence of a symptom would not necessarily mean that symptoms were the reason for the PSA test: a man might have had, say, erectile dysfunction, his doctor did not consider that related to prostate cancer, but agreed to give a PSA when the man requested it. Accordingly, we included an estimate for the proportion of patients who had an NHS-defined indicating symptom before a PSA, but the subsequent PSA test (therefore any resulting diagnosis) was not causally due to that symptom. In our base model, we set parameters so that the rate was 20% overall, that is, in most cases, the symptoms were the cause of the PSA. We further specified that this rate would vary by stage, that is, if a patient had symptom, then a PSA test and then was diagnosed with T3 or T4 disease, it was more likely that test was due to symptoms than if T1 or T2 disease was diagnosed. Hence, we used a rate 13% for T3/T4 and 27% for T1/T2, with both the absolute values, and the difference between stages, varied in sensitivity analyses. We separately evaluated a policy where PSA could not be given for cause in primary care. This was done by setting the probability of symptoms leading to a PSA test to a very low value, 0.1.

For T1/T2 cancers following asymptomatic PSA testing, the probability of overdiagnosis was taken to depend on the expected lead-time of cancer detection and life-expectancy. A statistical analysis^2^ of the European PSA screening randomized trial^3^ there estimated a lead time of about 10 years, a number we varied between 5 and 15 years in sensitivity analyses. We used a slightly more conservative assumption of 7 years, with 5 and 10 years used in a sensitivity analysis. Using the England and Wales life tables for 2017 – 2019, we calculated the probability that man would survive until the lead time if aged at the midpoint of each age decade (i.e. 45, 55, 65, 75 and 85 years). These life tables include death from prostate cancer, but this constitutes only a small fraction of deaths and so is unlikely to impact results. Although lead times and survival times have different distributions, the probability of surviving to the mean lead time is similar to the probability a lead time randomly selected from a distribution of lead times is less than a survival time randomly selected from a distribution of lead times. Moreover, any departures from this assumption would be incorporated in the sensitivity analysis modifying the lead time.

To calculate the annual overdiagnosis rate if screening were implemented at age 50 - 69, we used the overdiagnosis rates calculated for the current policy and made adjustments depending on age group. For men under 50 years of age, the number of patients overdiagnosed is very small and any adjustments would make not qualitatively change our findings. Under population-based screening men aged 50 – 69 years would receive an invitation to receive a PSA test, for which we assumed a base case compliance that 70% of invitees would attend (following evidence from breast and bowel screening in the UK)^4^, that 70% of invitees would attend^4^. Hence, the rate of screening-related overdiagnosed prostate cancer by age group was anticipated to increase by the difference between 70% and the current testing rate (excluding for cause) in each age group^5^ ^6^. For men aged 70 years or older, the base case scenario considered data from Lithuania^7^, one of the few countries to implement population-based screening, showing an ~80% reduction in the number of older men undergoing PSA testing after it was made available in a state-run program.

To estimate the number of PSA tests per year under current policy, we used data on age-group specific PSA screening rates^5^. To estimate the projected number of PSA tests under population-based screening, we used the following rates for each age group. For age <50 years, we assumed that rates would not change. For men aged 50 – 59 years, we followed European guidelines ^8^ that screening intervals are 5 years if PSA < 1 ng/ml, and 2 to 4 years if PSA is above 1 ng/ml, which we operationalized to every 4 years if PSA 1 – 2 and every 2 years if PSA 2 – 3 ng/ml. For men aged 60 – 69 years, we used a similar regimen, except there would be no additional screening were PSA < 1 ng/ml^9^. The proportion of the population in each PSA group was derived using normative data on PSA levels in a large unscreened Swedish cohort^10 11^. For men aged 70+ years, we assumed an 80% decrease in screening rates, as described above

Overdiagnosis and PSA testing rates under the current policy and under population-based screening were multiplied by estimates of the population size by age group, using population data from England^12^, to estimate the total number of overdiagnoses and PSA tests. The model was implemented in a Stata do file (see below).

We did not implement probabilistic sensitivity analyses in this study. Inputs to the model were derived either from population-based data (e.g. number of prostate cancer diagnoses in current practice) or were subject to sensitivity analyses across a much broader range than the confidence interval of a corresponding study. For instance, one input was the proportion of PSA tests that were “for cause”. Although the base case estimate (80%) was derived from a study, this was varied between 10% and 100% to reflect possible changes in policy and guidelines regarding the symptoms that would require a PSA test.

We triangulated our findings using an independent modeling strategy. This was derived using the cumulative incidence after screen detection from a screen using 15 year follow-up from the CAP trial^13^, where the excess to 15 years was estimated to 11.7% of screen-detected cancers, with the 95%CI (0.0% to 26.7%) used to define lower and higher overdiagnosis scenarios. To account for death from other causes, we used competing risks methods applying competing mortality rates based on English male mortality period rates^14^ and prostate cancer mortality rates by age (2018-20^15^]. The number of overdiagnosed cases by age group was estimated by multiplying the assumed number of asymptomatic prostate cancers detected by the corresponding estimated overdiagnosis rate. The base case for the number of asymptomatic cancers detected by age was from a 2018 study^1^ where n=871 men were diagnosed aged 70 years or older with asymptomatic prostate cancer, and n=982 aged 50-69 years. Change in overdiagnosis relative to this scenario was evaluated as a function of proportional increase in detection of men 50-69 and decrease in men 70 years or older. We then estimated the impact of increased asymptomatic cancer detection in men aged 50-69 years vs decrease detection in men aged 70 years or more. The code for this analysis can be found at <https://urldefense.com/v3/__https://github.com/brentnall/pca-overdx__;!!KVWo1iE!Ti8mr6ii88NvNKIR1W9jGxggOSKqRw2uot_oWukOZsGpfWHrE3kOsYA-VVpVkwvwmnsXsYBg-tVjjVHSGuHKiZ36$>.

**Stata do files**

There are two do files, one to compare overdiagnosis rates under current policy and under population-based screening and second to compare PSA testing rates under both scenarios. Both print out a table of results with cells delimited by “&”.

**Change in number of overdiagnoses**

**********************************************************************************

* Andrew Vickers January 10 2026

* calculate effect of implementing population based screening on overdiagnosis in UK

**********************************************************************************

quietly{

***ASSSUMPTION

*RATE OF SCREENING COMPLIANCE ith population based screening

* base assumption is to 70%

* UK FOBT program is about 67% https://www.gov.uk/government/publications/bowel-cancer-screening-annual-report-2023-to-2024/bowel-cancer-screening-standards-data-report-2023-24#bcsp-s02-uptake

*as is mammography https://www.england.nhs.uk/2024/01/new-breast-screening-figures-prompt-fresh-uptake-appeal/

local compliance_rate=.7

*store details of inputs is a local called “scenario” for later printing

local scenario="Compliance with population based screening: `compliance_rate'."

***ASSSUMPTION

*LEAD TIME

*use 7 (base assumption), 5, 10 or 15

local lead=7

local scenario="`scenario' Lead time: `lead' years."

***ASSSUMPTION: probability of for cause PSA

*if a man has a symptom before PSA that doesn't mean symptom cause PSA

* specify the probability that for a man with a symptom and then a PSA, the PSA was because of the symptom

* default is .8

*should be close to the probability of for cause biopsy below

local pifcpsa=.8

local scenario="`scenario' Probability that symptoms before PSA caused PSA `pifcpsa'."

***ASSSUMPTION

*REDUCTION IN SCREENING RATES IN OLDER MEN

* base scenario is represent 80% reduction (doi: 10.3390/jcm9123826.)

local reduction=.8

local scenario="`scenario' Reduction in screening in older men 0`reduction'."

***ASSSUMPTION

*PROPORTION OF OVERDIAGNOSIS IRRESPECTIVE OF LONGEVITY

* base case is 0%. alternative is 10% or 20%

local overdiagnosis_fixed=.1

local scenario="`scenario' Probability of overdiagnosis irrespective of life expectancy 0`overdiagnosis_fixed'."

***ASSSUMPTION: probability of for cause biopsy

*if a man has a symptom before PSA that doesn't mean symptom cause PSA

* specify the probability that for a man with a symptom and then a PSA, the PSA was because of the symptom

* default is .8

local pifc=.8

local scenario="`scenario' Probability that symptoms before biopsy caused PSA `pifc'."

***ASSSUMPTION differnece in for cause biopsy between T1T2 and T3T4

* We want to explore possibility that probability that a biopsy is for cause is different for T1T2 than T3T4

* say odds ratio of 2 for T3T4 vs. T1T2 vs. 1 in base scenario

local or =2

local scenario="`scenario' Odds ratio for T3vsT4 being for cause `or'."

*input data downloaded from https://nhsd-ndrs.shinyapps.io/rcrd/ for prostate cancer cases by age

*the data set gives data for 2019 for age groups <50, 50's, 60's, 70's, 80+

*create a data set of age groups and number of T1T2 and T3T4

clear

set obs 5

*create age group avariable

g agegroup="18-49"

replace agegroup="50-59" in 2

replace agegroup="60-69" in 3

replace agegroup="70-79" in 4

replace agegroup="80+" in 5

*input number of T1T2 and T3T4 into separate variables

*create and label the variables

g t1t2=.

g t3t4=.

label var t1t2 "Number of T1 and T2 cases per year by age group"

label var t3t4 "Number of T3 and T4 cases per year by age group"

*input the variables from a matrix where the numbers are stored

matrix input T1T2 =(359.0269775390625, 3343.35888671875, 7845.56201171875, 8513.986328125, 2301.658203125)

matrix input T3T4 =(141.9730224609375, 1823.64111328125, 5974.43798828125, 8655.013671875, 4997.341796875)

forvalues i=1/5{

replace t1t2=T1T2[1,`i'] in `i'

replace t3t4=T3T4[1,`i'] in `i'

}

*input probability of surviving to lead time , data from life tables

*Depends on user defined lead time

if `lead'==5{

matrix input LE=(0.987219979178038, 0.971720308827179, 0.930098706853037, 0.819791969238004, 0.521310437148245)

}

if `lead'==7{

matrix input LE=(0.980600906549838, 0.95668048060138, 0.894864123202306, 0.729382961948629, 0.356579774396768)

}

if `lead'==10{

matrix input LE=(0.968775311632956, 0.928390138746436, 0.8294060151066, 0.576563342877298, 0.167578291806409)

}

if `lead'==15{

matrix input LE=(0.941378645004122, 0.863494467503171, 0.679940390422085, 0.300568488319017, 0.0247674156386894)

}

*create a matrix to store current screening rates for 50- 59 and 60 - 69

*screening rate in under 50's not used so just given as 0

*(all data on screening rates from from table 1 of https://www.bmj.com/content/391/bmj-2024-083800.full)

*also see kiana-k-collins.shinyapps.io/bmj_shiny_app_v2

matrix input OSR=(0, .37 , .45)

*create a matrix to store current screening rates for cause for 50- 59 and 60 - 69

*screening rate in under 50's not used so just given as 0

*dta from Kiara collins

*https://pmc.ncbi.nlm.nih.gov/articles/PMC7805413/#table2

*first set of data from from https://pmc.ncbi.nlm.nih.gov/articles/PMC7805413/#table2

matrix input FCR=(0.295260489948538, 0.312117346310517, 0.363983891806933, 0.399382096238511, 0.380534670008354)

*now adjust for user defined probability of symptoms before PSA caused PSA

forvalues i=1/5{

local FCR`i'= FCR[1,`i']*`pifcpsa'

}

matrix input FCR=(`FCR1', `FCR2', `FCR3', `FCR4', `FCR5')

*create a matrix to store current rates of for cause cancers each age group <50, 50's, 60's, 70's, 80+

*derived from https://pubmed.ncbi.nlm.nih.gov/39401928/

matrix input FC=(0.702,0.764,0.788,0.818,0.873)

*adjust for user defined robability that symptoms before biopsy caused PSA

forvalues i=1/5{

local FC`i'= FC[1,`i']*`pifc'

}

matrix input FC=(`FC1', `FC2', `FC3', `FC4', `FC5')

* take into account probability that a biopsy is for cause is different for T1T2 than T3T4

*user defined odds ratio

forvalues i=1/5{

*get number t1t2s and t3t4s and total cancers into locals

local t1t2= t1t2[`i']

local t3t4= t3t4[`i']

local total =`t1t2'+`t3t4'

*updated odds in T3T4 is baseline odds (derived from probability) * OR

local odds=FC[1,`i']/ (1-FC[1,`i'])*`or'

*convert to a probability of for cause in T3T4

local pit3t4`i'=`odds'/(1+`odds')

*to calculate probability of for cause in T1T2...

* start by calculating absolute number of t3t4 that are for cause

local fct3t4`i'=`t3t4'*`pit3t4`i''

*calculate total number of for cause

local fc=FC[1,`i']*`total'

*subtract out to get t1t2 for cause

local fct1t2= `fc'-`fct3t4`i''

*probability of for cause in T1 t2

local pit1t2`i'=`fct1t2'/`t1t2'

}

matrix input FCT1T2=(`pit1t21', `pit1t22', `pit1t23', `pit1t24', `pit1t25')

matrix input FCT3T4=(`pit3t41', `pit3t42', `pit3t43', `pit3t44', `pit3t45')

**********************************************************************************

*now import life expectancy

*life expectancy is proportion surviving 10 years

*life expectancy is taken from england and wales life tables for 2017 - 2019 *https://www.ons.gov.uk/peoplepopulationandcommunity/birthsdeathsandmarriages/lifeexpectancies/datasets/nationallifetablesenglandandwalesreferencetables

*we used 10 year survival probability for age 45, 55, 65, 75, and 85 for the age groups respectively.

*england male population is taken from

*https://www.ethnicity-facts-figures.service.gov.uk/uk-population-by-ethnicity/demographics/age-groups/latest/

*breakdown by gender not given, so assume 50% male as reasonable approximation

*population is for england and wales, so take 95% of that to get England only

g le=.

label var le "Probabilty of surving n years, where n is from user input"

forvalues i=1(1)5{

replace le=LE[1,`i'] in `i'

}

*calculate current number of overdiagnosed cases

*approximated by the number of cases multiplied by proportion not surviving the mean lead time (give as le)

g current_overdiagnosed=t1t2*(1-le)*(1+`overdiagnosis_fixed')

label var current_overdiagnosed "current number of overdiagnosed cases"

**********************************************************************************

*calculate overdiagnosis rates under population based screening

*create a variable for number of T1T2 cases under population screening

g t1t2_screening=.

label var t1t2_screening "estimated number of T1T2 cases under population based screening"

*AGE GROUP <50

*. assume nothing changes for men < 50 (some good reason to get a PSA test if young)

replace t1t2_screening=t1t2 if age=="18-49"

*loop over age 50 - 70

forvalues i=2/3{

*get proportion currently diagnosed T1T2 cases for-cause PSA (e.g. DRE, urinary symmptoms, family history)

*remainder are screened

local fc=FCT1T2[1,`i']*t1t2[`i']

local sc=(1-FCT1T2[1,`i'])*t1t2[`i']

* calculate number of cases if screening increases

*put overall screening rate in a local (all PSA / population)

local ospi=(OSR[1,`i'])

*put for cause screening rates in a local (for cause PSA / all PSA)

local fcsr=(FCR[1,`i'])

*calculate proportion of population overall that is getting an asymptomatic PSA (asymptomatic PSA / population)

local psapi= `ospi' * (1- `fcsr')

*calculate proportion of population overall that is getting an for cause PSA (for cause PSA / population)

local fcpi= `ospi' * `fcsr'

*calculate proportion of population that would get asymptomatic PSA under screening

local targetpsa = `compliance_rate' - `fcpi'

*proportional increase in T1T2

local proportional_increase=`targetpsa'/`psapi'

*increase in cases

local sc=`sc' *`proportional_increase'

*total early cases under screening

replace t1t2_screening=`fc'+`sc' in `i'

}

*loop over age 70+

forvalues i=4/5{

*get proportion currently diagnosed cases for-cause PSA (e.g. DRE, urinary symmptoms, family history)

*remainder are screened

local fc=FCT1T2[1,`i']*t1t2[`i']

local sc=(1-FCT1T2[1,`i'])*t1t2[`i']

local sc=`sc'*(1-`reduction')

*total early cases under screening

replace t1t2_screening=`fc'+`sc' in `i'

}

**********************************************************************************

*compare overdiagnoses rates

*calculate overdiagnoses under population based screening

*approximated by the number of cases multiplied by proportion not living the mean lead time

* add in a certain proportion overdiagnosed irrespective

g screening_overdiagnosed=t1t2_screening*(1-le)*(1+`overdiagnosis_fixed')

label var screening_overdiagnosed "estimated number of overdiagnosed cases under population based screening"

*Calculate total number of cases and overdiagnoses under current practice and under screening

*Use capital letter to represent that these are total amounts

*calculate current number of early cases and overdiagnosis for early stage cancers and store in locals

egen CURRENT_OD=sum(current_overdiagnosed)

sum CURRENT_OD

local CURRENT_OD=r(mean)

egen CURRENT_CASEST1=sum(t1t2)

sum CURRENT_CASEST1

local CURRENT_CASEST1=r(mean)

egen CURRENT_CASEST2=sum(t3t4)

sum CURRENT_CASEST2

local CURRENT_CASEST2=r(mean)

*calculate projected number of early cases and overdiagnosis for early stage cancers under population based screening

egen PROJECTED_OD=sum(screen)

sum PROJECTED_OD

local PROJECTED_OD=r(mean)

local rr=round(`PROJECTED_OD'/`CURRENT_OD'*100,.1)

egen PROJECTED_CASES=sum(t1t2_screening)

sum PROJECTED_CASES

local PROJECTED_CASES=r(mean)

*drop temporary variables used to calculated totals

drop CURRENT_OD CURRENT_CASES* PROJECTED_OD PROJECTED_CASES

*save out results

sort age

**********************************************************************************

*Print out results

*create new row for total

set obs 6

replace age="Total" in 6

replace t1t2 = `CURRENT_CASEST1' in 6

replace t3t4 = `CURRENT_CASEST2' in 6

replace current=`CURRENT_OD' in 6

replace t1t2_screening=`PROJECTED_CASES' in 6

replace screening_overdiagnosed=`PROJECTED_OD' in 6

forvalues i=1(1)6{

if `i'==1{

noisily disp "Age group&Current T1T2&Current overdiagnoses&Projected T1T2&Projected overdiagnoses"

}

local a=age[`i']

noisily disp "`a'&" %-9.0fc t1t2[`i'] "&" %-9.0fc current[`i'] "&" %-9.0fc t1t2_screening[`i'] "&" %-9.0fc screening_overdiagnosed[`i']

}

}

disp "`scenario'"

disp "Relative proportion of overdiagnosis under population based screening " `rr' "%"

*save results to use to add to PSA results

save "Results OD.dta", replace

**Change in number of PSA tests**

**********************************************************************************

* Andrew Vickers October 10 2025

* calculate effect of implementing population based screening on PSA testing in UK

**********************************************************************************

*RATE OF SCREENING COMPLIANCE with population based screening

* base assumption is to 70%

* UK FOBT program is about 67% https://www.gov.uk/government/publications/bowel-cancer-screening-annual-report-2023-to-2024/bowel-cancer-screening-standards-data-report-2023-24#bcsp-s02-uptake

*as is mammography https://www.england.nhs.uk/2024/01/new-breast-screening-figures-prompt-fresh-uptake-appeal/

*https://www.england.nhs.uk/statistics/statistical-work-areas/screening/

local compliance_rate=.7

local scenario="Compliance with population based screening: `compliance_rate'."

*REDUCTION IN SCREENING RATES IN OLDER MEN

* base scenario is represent 80% reduction (doi: 10.3390/jcm9123826.)

local reduction=.8

local scenario="`scenario' Reduction in screening in older men 0`reduction'."

*start with UK male population

*population is absolute number

*uk male population is taken from

*https://www.ethnicity-facts-figures.service.gov.uk/uk-population-by-ethnicity/demographics/age-groups/latest/

*breakdown by gender not given, so assume 50% male as reasonable approximation

clear

set obs 6

g agegroup="80+"

replace agegroup="18-49" in 1

replace agegroup="50-59" in 2

replace agegroup="60-69" in 3

replace agegroup="70-79" in 4

g male_population=.

label var male_population "Male population of UK by age group"

matrix input MP= (3772250, 4076248, 3200360, 2574120, 1484913)

forvalues i=1(1)5{

replace male_population=MP[1,`i'] in `i'

}

*assume nelgible PSA tests in patients < 40

*ignore age less than 40 and adjust size of population

*add screening rates from https://kiana-k-collins.shinyapps.io/bmj_shiny_app_v2/

sort age

g current_screening_rate=.

replace current_screening_rate = 81 in 2

replace current_screening_rate = 162 in 3

replace current_screening_rate = 240 in 4

*for age group 80+, rates are given as 238 for 80-89 and 164 for 90+

*population over90 in UK from https://www.ons.gov.uk/peoplepopulationandcommunity/birthsdeathsandmarriages/ageing/bulletins/estimatesoftheveryoldincludingcentenarians/uk2002to2023

*constitutes about 1/3 of those >80

*take weighted average of screening rates in age 80-89 and 90+

replace current_screening_rate = (238*2+164)/3 in 5

*for men less than 40, take screening rate in those aged 40 - 49

replace current_screening_rate = 25 in 1

**********************************************************************************

*calculate current tests per year (remember rates are per 1000)

*assume nelgible PSA tests in patients < 40

g current_psa_tests=current_screening_rate*male_population/1000

**********************************************************************************

*estimate number of tets under population based screening

g projected_psa_tests=.

*for age 40 - 49, assume no change in rates

replace projected_psa_tests=current_psa_tests in 1

*for age 50 to 59, recommended intervals are:

* every 8 years for PSA < 1 (i.e. 0.2 per year) (European guidelines doi: 10.1016/j.eururo.2021.07.024.)

* every 2 to 4 years for PSA 1 - 3

* so say 4 years for PSA 1 - 2 (rate 0.25)

* and 2 years for PSA 2-3 (rate 0.5)

*quantiles (taken from Malmo data)

*60% <1	25% <1-2	15%>2 (includes pts with PSA > 3, so this an overestimate)

replace projected_psa_tests=((0.6*male_population*.2) + (0.25*male_population*.25) + (0.15*male_population*.5))*`compliance_rate' in 2

*for age 60 to 69, recommended intervals are:

* stop if PSA < 1

* every 2 to 4 years for PSA 1 - 3

* so say 4 years for PSA 1 - 2 (rate 0.25)

* and 2 years for PSA 2-3 (rate 0.5)

*quantiles (taken from Malmo data)

*50% <1	30% <1-2	20%>2 (includes pts with PSA > 3, so this an overestimate)

replace projected_psa_tests=(0.3*male_population*.25 +0.2*male_population*.5)*`compliance_rate' in 3

*for age 70+, stopping screening will reduce rates by 85% (doi: 10.3390/jcm9123826.)

replace projected_psa_tests=current_psa_tests*(1-`reduction') in 4/5

*Calculate total number of test current practice and under screening

*Use capital letter to represent that these are total amounts

*calculate current number of tests and store in local

egen CURRENT_PSA=sum(current_psa_tests)

sum CURRENT_PSA

local CURRENT_PSA=r(mean)

*calculate projected number of PSA tests under population based screening

egen PROJECTED_PSA=sum(projected_psa_tests)

sum PROJECTED_PSA

local PROJECTED_PSA=r(mean)

local rr=round(`PROJECTED_PSA'/`CURRENT_PSA'*100,.1)

*drop temporary variables used to calculated totals

drop CURRENT_PSA PROJECTED_PSA

**********************************************************************************

*Print out results

*create new row for total

set obs 6

replace age="Total" in 6

replace current_psa_tests = `CURRENT_PSA' in 6

replace projected_psa_tests=`PROJECTED_PSA' in 6

l

disp "`Scenario'"

disp "Relative proportion of PSA tests under population based screening " `rr' "%"

sort age

merge 1:1 age using "Results OD.dta"

drop _m

l

forvalues i=1(1)6{

if `i'==1{

disp "Age group&Current PSA tests&Current T1T2&Current overdiagnoses&Projected PSA tests&Projected T1T2&Projected overdiagnoses"

}

local a=age[`i']

disp "`a'&" %-10.0fc (current_psa_tests[`i']) "&" %-9.0fc t1t2[`i'] "&" %-9.0fc current_overdiag[`i'] "&"%-10.0fc (projected_psa_tests[`i']) "&" %-9.0fc t1t2_screening[`i'] "&" %-9.0fc screening_overdiagnosed[`i']

}
